# The COVID-19 Incarceration Model: a tool for corrections staff to analyze outbreaks of COVID-19

**DOI:** 10.1101/2021.02.18.21252032

**Authors:** Jisoo A. Kwon, Neil A. Bretaña, Luke Grant, Jennifer Galouzis, Wendy Hoey, James Blogg, Andrew R. Lloyd, Richard T. Gray

**Author notes:** **Correspondence to:** Dr Jisoo Amy Kwon, Surveillance Evaluation and Research Program, The Kirby Institute, UNSW Sydney, Sydney NSW 2052, Australia.

## Abstract

Correctional facilities are at high risk of COVID-19 outbreaks due to the inevitable close contacts in the environment. Such facilities are a high priority in the public health response to the epidemic. We developed a user-friendly Excel spreadsheet model (building on the previously developed Recidiviz model) to analyze COVID-19 outbreaks in correctional facilities and the potential impact of prevention strategies - the COVID-19 Incarceration Model. The model requires limited inputs and can be used by non-modelers. The impact of a COVID-19 outbreak and mitigation strategies is illustrated for an example prison setting.

## Introduction

Substantial outbreaks of COVID-19 illness have occurred across the world within correctional facilities including prisons, jails, and detention centers (1-3). Such facilities, termed here as “prisons”, are at particular risk of COVID-19 outbreaks due to the dynamic interactions with the wider community and the largely unavoidable close contacts that occur within these settings. Inmates, correctional and healthcare staff, and visitors are all at risk of infection if an outbreak becomes established. Inmates may also be particularly vulnerable to COVID-19 illness, as well as to the accompanying morbidity and mortality, due to their high prevalence of underlying health conditions. In the United States, between 74-98% of infection rates among the prisoners in correctional facilities in Ohio, California, and Louisiana (4). Prisons are therefore a high priority setting in the public health response to the COVID-19 pandemic, and appropriate prevention and mitigation strategies need to be developed by correctional departments as part of the wider response (3).

Mathematical models are useful tools for understanding infectious disease outbreaks, including explaining epidemiological patterns, evaluating the population-level impact of public health control programs, and forecasting epidemic trajectories (5, 6). Models can also be used to guide organizational responses by evaluating the potential impact of interventions to prevent or mitigate an outbreak of COVID-19 in prison settings (7-9). Traditionally, the development of epidemiological models is done by specialist researchers with the appropriate software coding skills, epidemiological expertise, and mathematical expertise. The development of many mathematical models requires significant time and resources followed by adaptation to local data inputs, hence prison settings are unlikely to be able to develop a model, acquire timely epidemiological information, and then apply the model to inform responses relevant to the local setting.

We developed a simple modelling tool within Microsoft Excel (Redmond, WA) for simulation of outbreaks of COVID-19 within prisons, and the potential impact of control interventions. Our model was originally designed to model potential outbreaks within the state prisons service in the New South Wales, Australia and to assess the impact of intervention programs, including personal protective equipment (PPE), isolation, quarantine, thermal testing, reducing prisoner population, and reducing visitors. However, the finalized model can be customized to the unique behavioral and epidemiological context of any prison or other closed setting. The model is sufficiently user-friendly to ensure that untrained correctional staff can perform their own analyses.

The COVID-19 Incarceration Model incorporates virus transmission between inmates, correctional staff, healthcare staff, and visitors. It allows designation of the prevalence of vulnerabilities to severe COVID-19 in the population, varied numbers of close contacts, and the daily intake and release of inmates. We developed the model by modifying and expanding the scope of an existing Excel spreadsheet model developed by the non-profit organization Recidiviz (10), to capture additional complex features of prisons, a broad range of prison specific interventions for control of COVID-19 spread, and the mixing patterns between inmates and staff. It incorporates the latest international evidence on COVID-19 transmission and disease progression (Table 1).

**Table 1:**
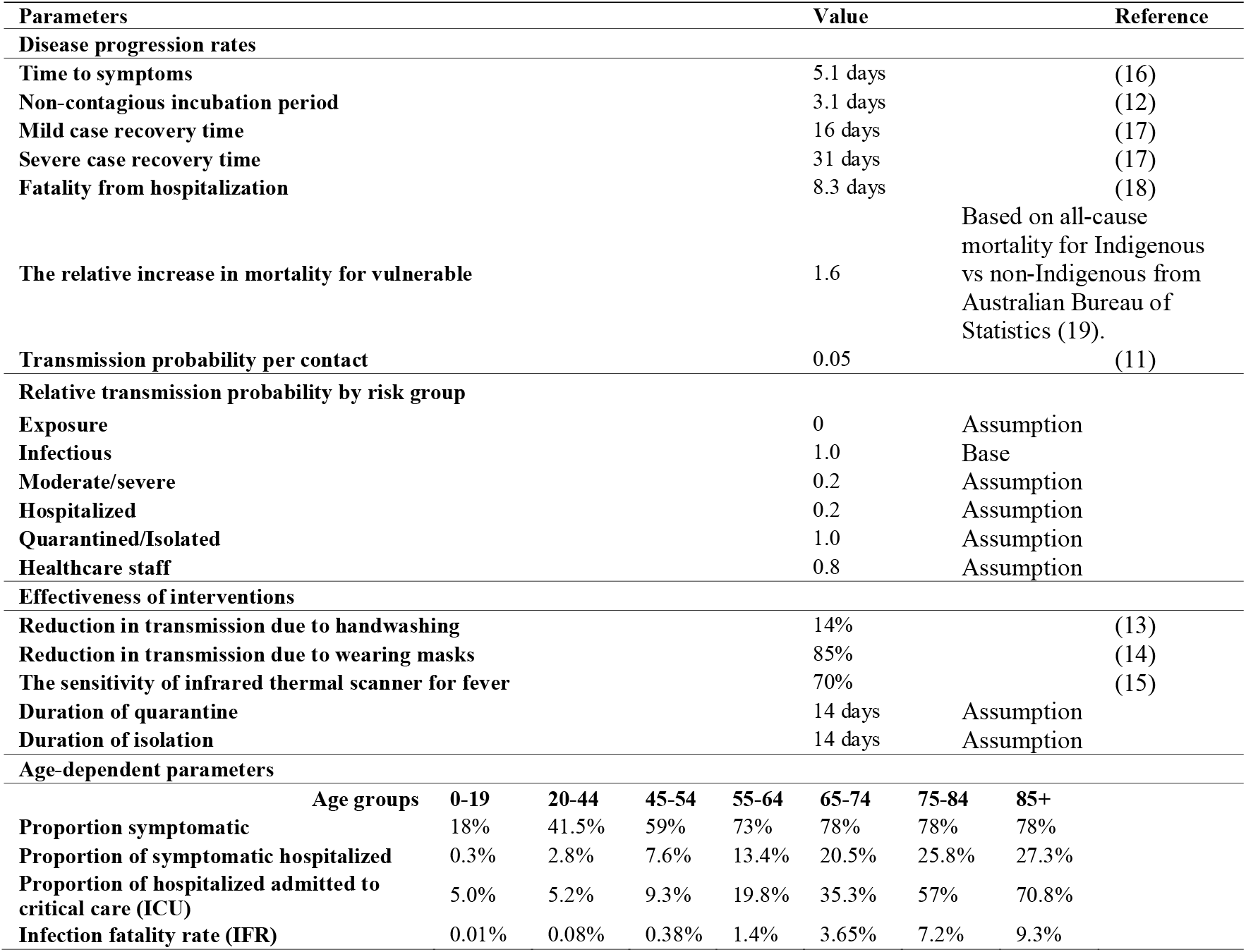
Model inputs and parameter estimates

The Model ensures flexibility by allowing users to define and assess different outbreak scenarios and combinations of targeted control strategies to illustrate epidemic patterns and the effect of prevention or mitigation programs. The model is easily adopted for application to different prison settings. In this manuscript, we will demonstrate how the model works and its use for assessing the potential outcomes of a COVID-19 outbreak in a generic prison setting and the impact of associated interventions.

## Method

To track an outbreak of COVID-19 in a prison setting and assess the potential impact of prevention strategies and interventions, we developed a simple compartmental deterministic model of COVID-19 transmission and disease progression within a Microsoft Excel spreadsheet (Redmond, WA). The model substantial extends the previously developed Recidiviz model to include key features of COVID-19 transmission, detailed demographics characteristics of inmates and prison staff, and more complex intervention strategies. Our Excel spreadsheet model is publicly available under an open-access license (GNU General Public License, Version 3) via https://github.com/The-Kirby-Institute/covid19-closed-pop-models. The version used to generate the results in this manuscript is provided as supplementary material along with a user manual.

### Model structure

The model splits the prison population into the following compartments representing the overall number of inmates and staff who are: susceptible; exposed; infectious, with mild illness, with severe illness, hospitalized; and recovered (Figure 1). It then tracks the number of people in each compartment over time and calculates the number of deaths and new infections separately. Inmates are grouped by age in the model along with the proportion in each age group ‘vulnerable’ to severe COVID-19. The inmate population can be treated as a whole or split into seven-year age cohorts (0-19, 20-44, 45-54, 55-64, 65-74, 75-84, 85+). The model is implemented in a framework that updates the number of people in each compartment each day over a 120-day period. This is done using what is known as difference equations which are incorporated into the model and calculate the number of people who enter and leave each compartment each day to update the number for the next day. These calculations are done ‘internally’ using formulas within the model spreadsheet. For the infected population groups, COVID-19 disease progression is given by daily rates with assumptions based on up-to-date data from international settings of COVID-19 infection. This includes age-specific values for the development of symptoms, severe disease, hospitalization, and death (Table 1).

**Figure 1:**
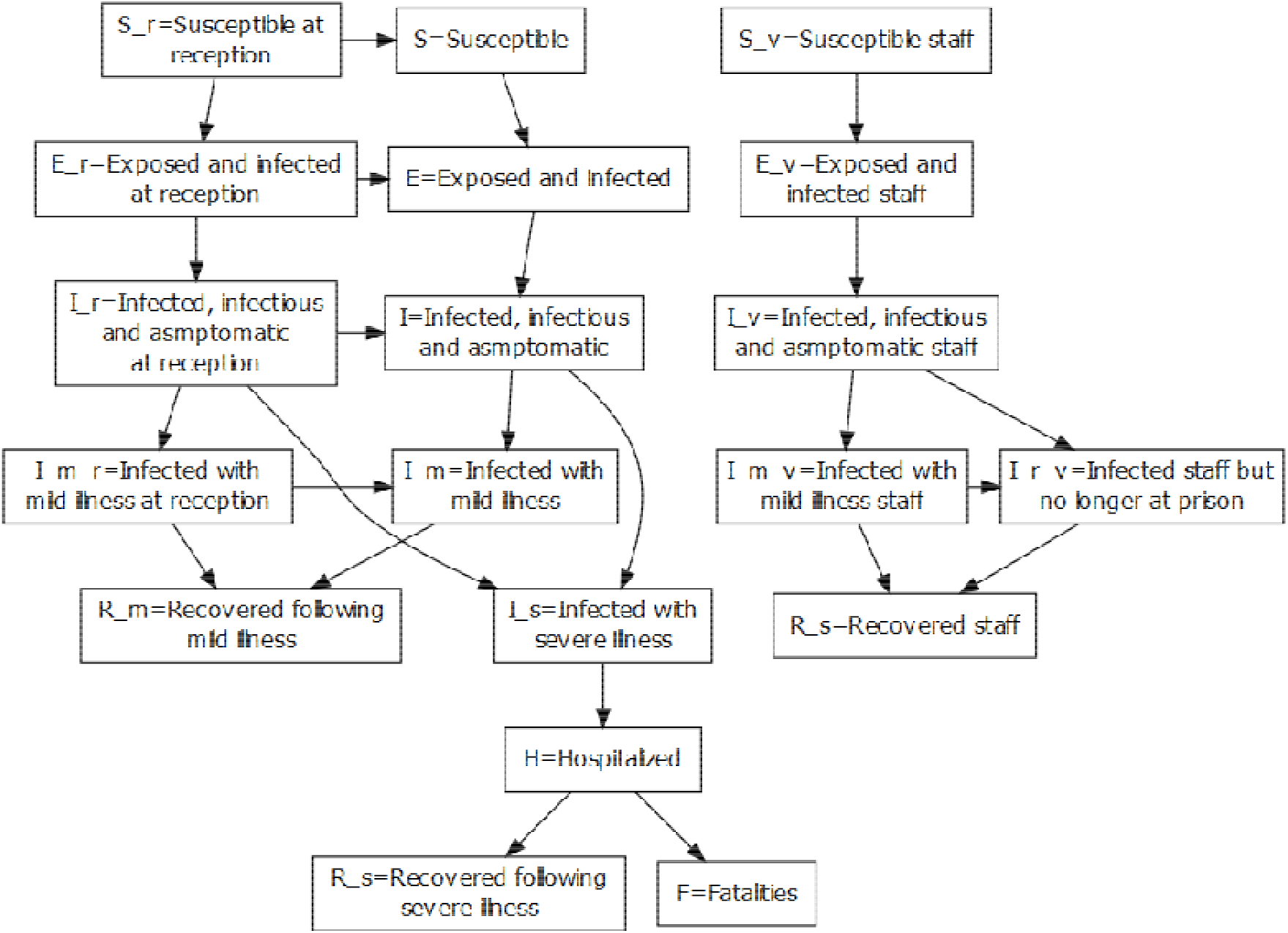
Schematic diagram of the COVID-19 incarceration model showing disease progression.

The model is specifically designed for each prison setting with the daily number of inmates coming into the prison (either from the community or via transfer from another prison setting), the daily number of visitors, and staff working at the site described. As inmates enter the prison, they are allocated to age groups based on the age distribution of current inmates. The discharge of inmates is also described (but the model does not track where they go), with the number of inmates who are infected when they are discharged recorded. However, within the model, inmates are not released while they are symptomatic. Staff are assumed to attend the prison site every day, while family visitors only attend the site once.

The COVID-19 transmission from infected to susceptible people is via close contact (defined to be less than 1.5 meters for longer than 15 minutes)) with a probability of transmission per contact determining the number of contacts becoming infected (11). This transmission probability varies to reflect susceptibility by age and the use of PPE (including mask use, hand washing, and hygiene measures). We also incorporated differences in transmission risk by disease stage (Table 1) as the level of viral shedding changes during the course of infection (with infectiousness highest during the pre-clinical and early symptomatic stages) (12). Hospitalized patients and healthcare workers are assumed to have access to sufficient PPE in the model (Table 1). In the model the number of contacts is specified by the user for each population group with contacts between inmates and staff across the prison being completely random (known as homogeneous mixing). The model can initiate outbreaks via inmate or staff or family visitors on a specific date.

### Interventions incorporated into the model

Several interventions to reduce the risk of COVID-19 transmission are incorporated into the model (Figure 2). These interventions are specified by changing model inputs/parameters, the coverage or number/proportion of people exposed to the intervention, and the time an intervention is introduced relative to the start of an outbreak.

**Figure 2.**
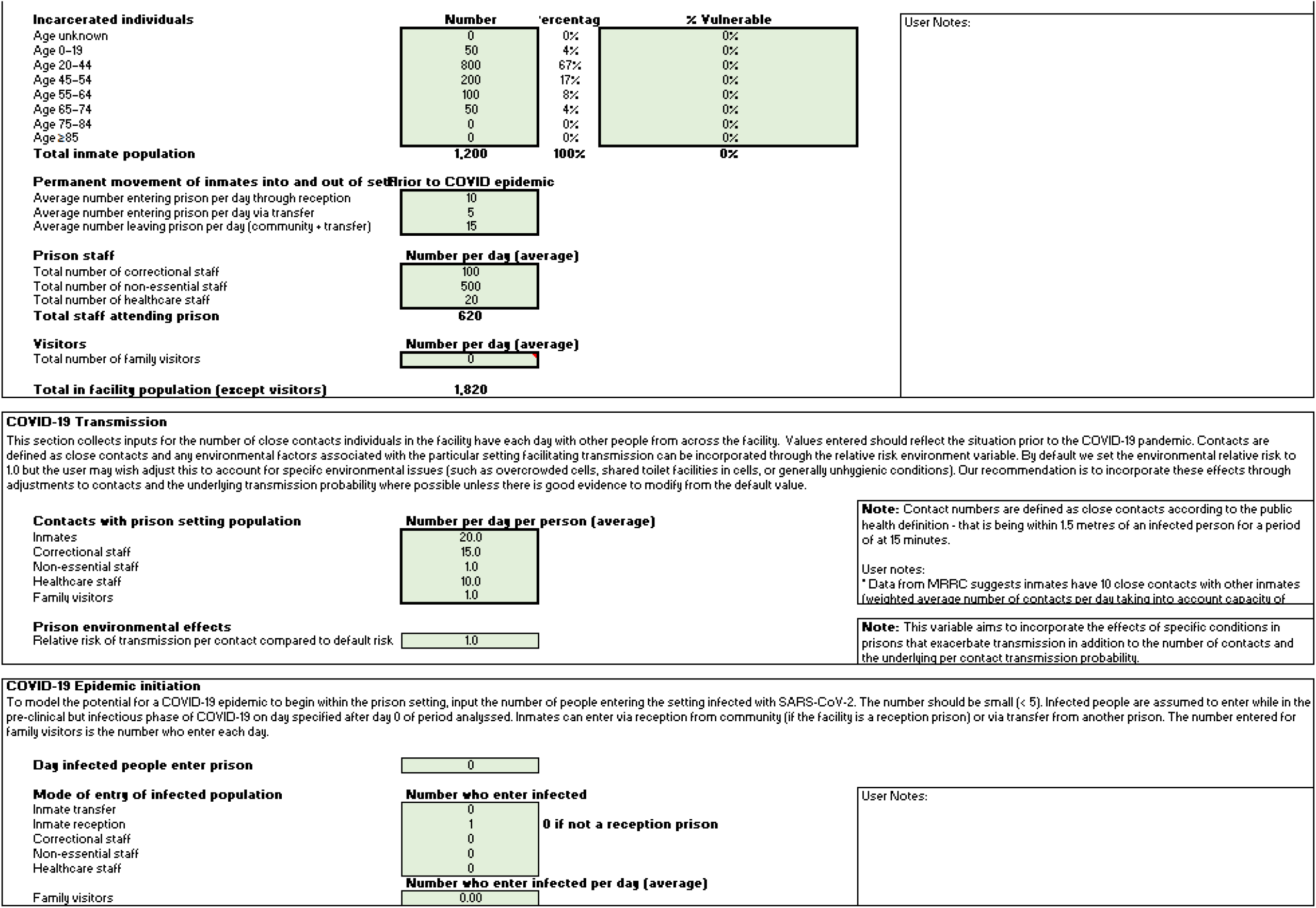
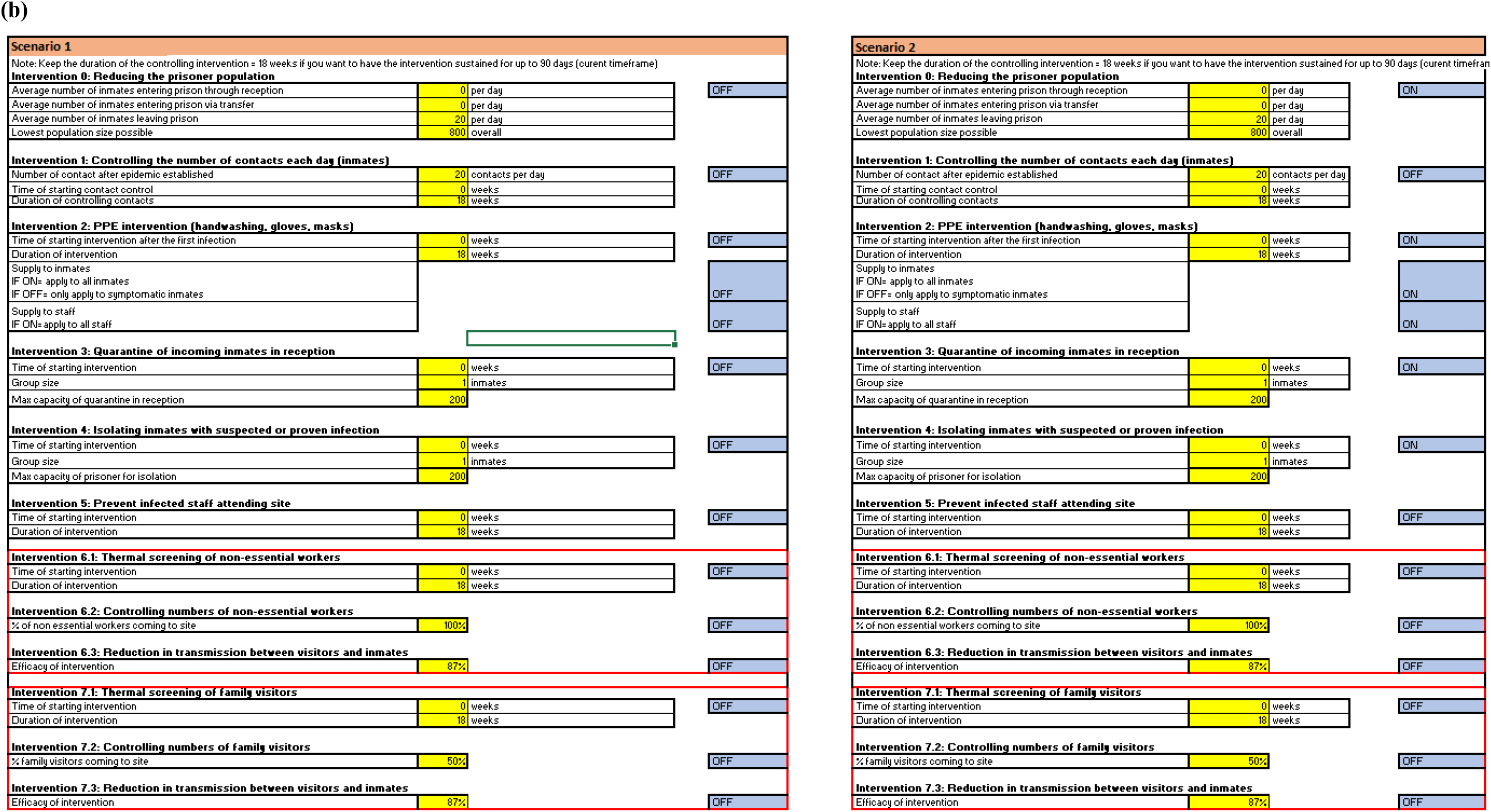
Snapshot of the main ‘input’ sheet: (a) storing information on potential relevance to the prison setting being analyzed; (b) specific interventions turned on and off (using toggle cells shaded in blue) or a combination of interventions to be compared. Numbers are for example only.

The specific interventions incorporated into the model are described below with an explanation of how they are implemented in the model:

- *Intervention 0 -Deferral/early release:* where the number of inmates in the prison is reduced over time to reduce the population at risk of COVID-19 infection. Such a reduction could reflect a policy of deferral of sentencing, or early release of low-risk inmates. Within the model, the number of inmates entering and leaving prison each day can be specified by the user to reflect the linear change in population size over time. For this intervention, a minimum population that can be reached can be specified to reflect the existence of inmates who cannot be released.
- *Intervention 1 - Reduction in contacts:* where the overall average number of contacts inmates have each day is reduced. This intervention groups together the effects of measures that promote social distancing (e.g. reductions in time out of cell, prevention of congregations, cancelling of work, group counselling or training). Within the model, the effects of these interventions are simply captured by changing the value of the number of contacts for inmates which reduces the probability of infection.
- *Intervention 2 - Quarantine at reception:* the model can reflect the effects of quarantine of inmates entering prison from the community to prevent spread from those who enter infected. In the model, we assume quarantine lasts for 14 days with incoming inmates quarantined by themselves within single cells or in groups (to reflect logistical constraints that are likely be present within a prison). The user specifies the size of the quarantine groups (1 or above), the day post-outbreak that quarantine begins, and a maximum number of people that can be quarantined at any one time, again to enable logistical and structural constraints to be considered. When the maximum number of people in quarantine is reached the model assumes any additional new inmates go into the prison without quarantine. While in quarantine, inmates can still have contacts and transmit to other people but at a much lower rate. The number of contacts a quarantined individual has depends on the size of the cohort. Finally, the compartmental structure of the model means that even though people are in quarantine for 14 days, which is an average, and infected people can leave quarantine early and potentially cause an outbreak. This “leakage” of infection is realistic given quarantined inmates will still have to interact with staff and others to receive meals, access sanitation, and receive healthcare.
- *Intervention 3 - Isolation of inmates:* where inmates already in the general prison population who are diagnosed, or those suspected to be infected (due to symptoms or being a close contact of an infected person) are isolated from the rest of the inmate population. In the model, this intervention is implemented and specified in the same way as for quarantine on entry within inmates placed in isolation for 14 days individually or within groups up to a maximum number.
- *Intervention 4 – Provision of PPE to staff/inmates:* where the effects of providing masks, gloves and handwashing materials to inmates and facility staff are modelled. Within the model, the effects of these measures are captured by a combined relative reduction in the transmission probability using data from published literature (13, 14) (Table 1). Available efficacy estimates were generally obtained from studies in health care settings, which are likely to have much better infection control standards than prisons. We, therefore, assumed the efficacy of these interventions to be half that of a healthcare setting. This results in PPE reducing the probability of transmission during close contact by a default 56% in the model, but this can be changed by the user. Healthcare workers are assumed to always have sufficient disposable PPE when seeing patients. The model allows PPE to also be available to other staff only, and to all inmates.
- Intervention 5 – *Prevent infected staff attending site:* this intervention ensures staff who are symptomatic isolate themselves and not attend the site while infectious. We assume staff are still attending the prison site if they are in the susceptible or exposed stages. When this intervention is in place staff who are symptomatic are counted in a separate model compartment until they recover and cannot transmit the virus to other staff or inmates in the prison.
- *Intervention 6 - Thermal screening of non-essential staff and family visitors:* this intervention captures the effects of visitors to the site being screened for fever upon entry. We assume the sensitivity of screening in the model is 70% (15) and this reduces the probability of transmission from a visitors to other people at the prison setting by a corresponding 70%, but this can be changed by the user.
- *Intervention 7 - Reductions in the number of non-essential staff and family visitors:* this intervention aims to capture the potential effects of stopping visitors (either temporary workers or family visitors) coming to the prison. The user enters the percentage reduction in the number of non-essential staff and family visitors attending the prison each day. The model assumes the same number of visitors enter the prison each day and have a specified number of contacts. We assume the number of contacts is small which means that an outbreak is not certain even if an infected visitor enters the prison each day—rather outbreaks will occur probabilistically or stochastically. Our model is not designed to explicitly capture this type of effect, but it can be reflected by estimating the risk and the expected time until an outbreak occurs if infected visitors enter the prison each day.

Within the spreadsheet, the scenario input parameters can be entered by the user and turned on or off via the ‘Inputs’ sheet to capture the effects of each intervention individually or in combination.

### Model parameters

Key factors or model parameters describing the COVID-19 transmission probability, disease progression and recovery rates, the probabilities of developing symptoms, the relative effectiveness of interventions on transmission, the relative effects of age on hospitalization, need for intensive care, and mortality are included in the model. These parameters are set at constant default values based on available literature as described in Table 1. The values are stored on the ‘Variables’ sheet in the model spreadsheet with notes and references providing justifications for these values, which can be updated by the user if required, to better reflect the characteristics of the setting modelled or updated data.

### Date requirements

To use the model, a minimal number of inputs are required but more comprehensive input data will improve the simulation of an outbreak. To run the model the minimum data required include: an estimate for the inmate population size; the average number of contacts per inmate per day; and the number of correctional staff, healthcare staff, non-essential staff, and family visitors. The inmate population can be split into age groups to better reflect the risks of severe COVID-19 if there is sufficient data. In addition, the percentage of inmates that are particularly vulnerable to severe COVID-19 can be entered, but this is not essential for the model to run. The number of inmates that enter and leave the prison each day also needs to be entered to understand the impact of changes in prisoner population size on an outbreak; otherwise the model assumes the prison is a closed population. To assess the risk of a visitor initiating an outbreak. the average number of visitors to the facility each day needs to be entered.

Besides the inmate population size, the number of contacts per day is a key input into the model as it affects the size and speed of the outbreak and the relative effect of interventions. Information on the number of contacts per person each day is not often available, but can be informed by: the average number of inmates in each cell, area and pod in each prison; the size of work, training, or exercise groups; the number of patients each healthcare staff see each day; the number of correctional staff working each shift and attending change-over meetings; and finally by surveys of staff and inmates aimed at estimating the number of contacts. An alternative if contact information is not available is to “calibrate” the number of contacts so that the model simulation outputs match the expected growth rate and size of an outbreak based on case data from similar prisons, or even the modelled prison itself if an outbreak has already occurred.

### Outputs

Once all the inputs are entered, the model produces simulated outbreaks and records daily numbers of new infections, the number of people living with COVID-19 while in each disease stage (asymptomatic, mild illness, moderate-severe illness, in hospital, and recovered) for each inmate and staff population group for 120 days. From these daily estimates, the model outputs the cumulative number of new cases and deaths at 7, 30, 90, and 120 days for inmates, and for the staff populations. For staff, the model provides daily estimates for the number infected and requiring time off work.

While we assumed infected inmates stay in the prison until recovered, inmates who are exposed or asymptomatic/pre-clinical can leave the prison if their length of stay finishes, which could lead to leakage of COVID-19 outbreaks to the community or upon transfer to a different prison. We, therefore, tracked the number of infected inmates (asymptomatic/pre-clinical) who leave the prison each day in the model.

### Model spreadsheet structure and its use

The model spreadsheet consists of six separate sheets which are used to enter model inputs (‘Inputs” sheet), show model outputs (‘Outputs’ sheet), contain the underlying model equations and perform the scenario calculations (‘time-dependent_scenario1’ & (‘time-dependent_scenario2’, and ‘Calculation_scenario1’ & ‘Calculation_scenario2’), store the default model parameters (‘Variables’ sheet), and information about the model including the software license information (‘Information’ sheet). The two most important sheets for using the model are the ‘Inputs’ and ‘Outputs’ sheets. The ‘Variables’ and ‘Information’ sheets are primarily for providing information, although model parameters can be changed on the ‘Variables’ sheet if required. The scenario calculation sheets are read-only and can be essentially ignored by users (although advanced users can find additional results for each scenario). More information can be found in the Supplementary manual.

The ‘Inputs’ sheet is where users enter model inputs. This sheet is split into two sections where the top section is used for entering the input data and information for the specific prison being analyzed (Figure 2 (a)). The bottom section is for specifying the scenarios that are simulated by the model (Figure 2 (b)). The model can run two outbreak scenarios simultaneously (‘Scenario 1’ and ‘Scenario 2’). Intervention inputs can be specified to assess the potential impact of various combinations of interventions on COVID-19 transmission. In each scenario, specific interventions are turned on and off to enable the impact of a single intervention (using toggle cells shaded in blue, ‘Input’ spreadsheet, Figure 2 (b)) or a combination of interventions to be compared. This includes a “no-response” scenario where all interventions are turned off, allowing the model to track the trajectory of an unmitigated outbreak where all parameters are set to their values prior to the appearance of the outbreak. The ‘Output’ sheet shows the main results and graphs for scenario 1 and scenario 2. The charts and tables on this sheet show the model outcomes of the two scenarios specified on the ‘Input’ sheet. The results include a number of new infections, asymptomatic, mild illness, moderate-severe illness, in hospital, recovered, and deaths for both inmates and staff.

### Demonstration model and example analyses

To demonstrate the use of the model and illustrate the potential impact of interventions to prevent and/or mitigate an outbreak of COVID-19 within a prison setting, we developed a model for an example prison with 1,200 inmates, 100 correctional staff, 20 healthcare staff, and with no visitors. For this model, we assumed inmates and staff had 20 and 15 close contacts on average per day - as shown in Figure 2 (a).

In this model we ran two scenarios, a ‘No-response’ scenario (Scenario 1) where no interventions are in place compared to a scenario where there is a reduction in population size (of net 20 inmates per day down to 67% of the initial population), PPE is being used by all staff and inmates, quarantine of new inmates on an entry for 14 days individually up to a total of 200, isolation of symptomatic and diagnosed inmates for 14 days individually up to a total of 200 are implemented one at a time and simultaneously in combination (see Figure 2 (b)). Under these scenarios, we project the potential epidemic of SARS-CoV-2 within inmates and staff if one infected inmate entered the prison from the community on day zero.

## Results

The model shows that if no response were put in place with an outbreak (Scenario 1) in the example prison then almost 100% of inmates would become infected over 120 days while in prison (assuming entry and discharge of inmates continues) (Figure 3 (a)). Over this period, all staff could also become infected. The peak prevalence of active infection could reach 80% within inmates on day 30. At the peak of prevalence, almost 80% of correctional and 70% of healthcare staff would also be infected and unable to attend work at the prison (Figure 3 (b)).

**Figure 3.**
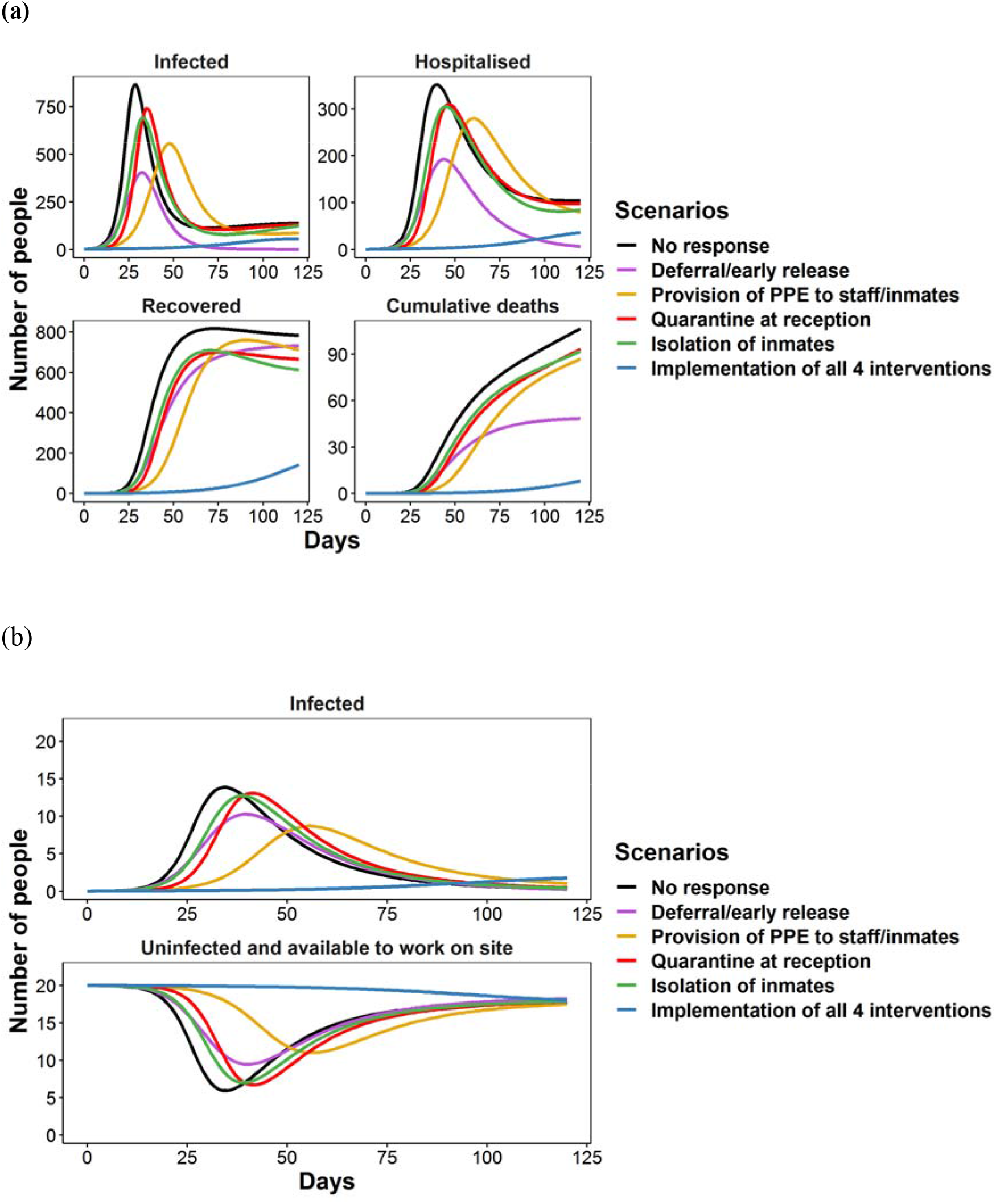
Change in the number of inmates infected, hospitalized, recovered and the number who have died during an outbreak in the reception prison (receiving new inmates from the community) under the no-response and intervention scenarios for: (a) inmates; and (b) all staff (correctional and healthcare staff). Each intervention is applied separately and in combination.

Each control intervention, when implemented separately, resulted in a reduced and delayed peak in infection (Figure 3 (a)). PPE had the biggest impact in delaying the peak from day 30 to day 48 with 20% reduction in the cumulative number of infections, followed by quarantine of inmates on entry (peak on day 35), isolation of inmates (peak on day 33), and reducing the prison population size (peak on day 32). In combination, the interventions could prevent an outbreak from occurring.

The model also shows that if there is no response to an outbreak in the prison, then an estimated maximum number of 350 inmates would require hospital beds, and a peak of 60 ICU beds on day 40. Each intervention, when implemented separately, would reduce the number of hospital and ICU beds required. In combination, the intervention resulted in a 90% reduction of hospital and ICU bed needs in the facility, respectively.

## Conclusions

We developed a simple and widely applicable Excel spreadsheet model of COVID-19 transmission within correctional facilities. This user-friendly model was designed to be used by correctional and prisoner health staff to estimate the potential impact of COVID-19 on inmates, and on staff; as well as the effects of prevention and mitigation strategies. In this manuscript, we present the results from a demonstration prison to show how the model works and to illustrate the potential impact of COVID-19 on inmates and staff and the effect of plausible interventions.

Our model shows that almost 100% of inmates and staff would become infected over 120 days if there is no response to an outbreak in the example prison. The combination of all plausible interventions (Figure 3 (a)) including widespread of PPE use, isolation of inmates, quarantine of new inmates on entry, and reducing the prisoner population size, can prevent an outbreak from occurring.

It is important to note that the Excel spreadsheet model we have developed is a relatively simple model and does not capture all the complexities of interactions within a prison, or the specific interactions between individuals. This means it may overestimate the magnitude of an outbreak in a prison where the internal structure includes multiple wings and yards that can be isolated from each other in the event of an outbreak. Our model is a deterministic model which means it does not capture probabilistic effects when the infection numbers are small, nor the risk of an outbreak initiating due to the presence of an infected individual. Rather the model assumes an outbreak will occur once an infected individual enters and shows the resulting trajectory of such an outbreak. The model describes the movement between quarantine and isolation and the general inmate population as an average rate equal to the inverse of the quarantine/isolation period. This means there can be a slow release of infected individuals from quarantine/isolation in the model sparking off an outbreak earlier than what might be expected. Depending on the intervention parameters this is shown as a delayed trajectory with a slightly lower peak. However, inmates in quarantine/isolation can still have some interaction with staff and exposed individuals may be released at the end of the quarantine/isolation period meaning this slow spread of infection from quarantine/isolation is not unrealistic. Finally, this simple model does not describe the impact of varied testing strategies for COVID-19.

Many of these limitations can be addressed by developing a more detailed and sophisticated model. Nevertheless, this user-friendly model was specifically developed to provide an accessible tool to guide prevention strategies to quickly assess the impact of interventions to mitigate outbreaks in the prison setting. This model can be used by non-modelers and can be readily applied to other closed population settings.

## Data Availability

The model is publicly available under an open-access license via https://github.com/The-Kirby-Institute/covid19-closed-pop-models. The version used to generate the results in this manuscript is provided as supplementary material along with a user manual.

## Acknowledgements

This study was funded from Corrective Services NSW Australia. The Kirby Institute is funded by the Australian Government Department of Health and is affiliated with the Faculty of Medicine, UNSW Sydney. The views expressed in this publication do not necessarily represent the position of the Australian Government. JAK produced results, interpretation of findings, and wrote the manuscript; NAB provided interpretation of findings and manuscript writing; ARL led the study, providing oversight in the design, implementation, interpretation of findings, and manuscript writing; LG led the study, providing oversight in the design, implementation, interpretation of findings, and manuscript writing; JG, WH, and JB assisted in data collection and editing the manuscript; RTG led the study, providing oversight in the design, implementation, interpretation of findings, and manuscript writing. We acknowledge Nicola Archer-Faux from Department of Communities and Justice, Kimberley Conlan from Corrective Services NSW, Joshua Taylor from NSW health, Colette Mcgrath from Justice Health Forensic Mental Health Network NSW, Kevin Corcoran from Corrective Services NSW, Andrew Warren and Justine Kunz from the Recidiviz team, and David Boettiger from the Kirby Institute, UNSW for their input.

