## Supplementary material for "The COVID-19 Incarceration Model: a tool for corrections staff to analyze outbreaks of COVID-19": Supplementary-COVID-Incarceration-Manual.docx

**1. Introduction to COVID-19 Incarceration Model**

**COVID-19 Incarceration Model**

The purpose of this interactive modelling tool is to project the potential impact of COVID-19 transmission within prison settings. It is developed in Microsoft Excel to allow for general access without dedicated software. The tool is based on the model developed by Recidiviz ([www.recidiviz.org](http://www.recidiviz.org)). Our tool assesses the impact of different strategies (scenarios) in preventing or mitigating COVID-19 outbreaks within prison settings. It was initially used for prisons in the Australian state of NSW but has been flexibly designed for prison settings in general. This guide provides an overview of how to use the COVID-19 Incarceration Model to assess the potential impact of COVID-19 outbreaks in prison settings.

The model spreadsheet contains six main worksheets: Inputs, Output, Variables, time-dependent_Scenraio1, time-dependent_Scenraio2, Calculation_Sceanrio1 Calculation_Sceanrio2, and Information. The *Inputs* sheet is where users enter specific prison setting data and specify scenarios and input values for the scenarios. The *Output* sheet is where the main outcomes of the model for each scenario are shown. The *Variable*s sheet stores the underlying diseases progression and transmission parameters obtained from publically available literature. The *Information* sheet provides general information about the project, a compartmental model diagram, and license information. The other sheets contain the model calculations and input parameter adjustment calculations. Users should primarily use the *Inputs and Outputs* sheets.

The model has been made publicly available for transparency and replication purposes and in the hope that it will be useful under the terms and conditions of the GNU General Public License. We take no responsibility for results generated with the spreadsheet and their interpretation but are happy to assist with its use and application.

**2. Input data: viewing and editing data**

The user enters inputs primarily on the *Inputs* sheet in the green shaded cells and with scenario inputs entered in yellow shaded cells. The input parameters for the model are on the *Variables* sheet. These are based on the available evidence but can be changed by the user if desired. Descriptions of each variable contained in these sheets are explained as follows:

**Regional Context (*Inputs* sheet)**

This section is for storing information on potential relevance to the prison setting being analysed. The only numbers used in the analysis are the number of hospital and ICU beds available.

**Setting Demographics (*Inputs* sheet)**

This section of the input sheet is for entering general population demographics of the prison setting being analysed.

***Population size:*** Estimated population size of inmates by age group.

***Vulnerable (%):*** Proportions of the vulnerable population by age group.

***Permanent movement of inmates into and out of setting (prior to COVID epidemic):*** Baseline population movement prior to COVID-19 epidemic. This is the average number of inmates entering and leaving the prison setting permanently each day. Inmates can enter the prison through reception or from another centre.

***Number of staff:*** Number of correctional, non-essential, and healthcare staff attending prison per day on average. Non-essential staff includes volunteers, contractors, etc who are not essential for the day-to-day running of a prison.

***Number of family visitors***: Number of family visitors of inmates per day on average.

**COVID-19 Transmission (*Inputs* sheet)**

***Average number of contacts:*** The average number of close contacts per person per day for each population group. The model assumes a homogeneous mixing of age-groups and populations across the prison setting. These parameters (along with the transmission per contact input on the *Variables* sheet) define the risk of COVID transmission from one inmate/visitor/staff to another inmate/visitor/staff. We consider a contact to be a close contact where transmission is likely to occur in line with the public health definition for contact tracing (e.g. 15 minutes within 1.5 metres). The average number of contacts needs to be estimated as best as possible using data on the number of people in each cell (multi-cell and single-cell), yard capacity, or the number of people interacting during daily activities (including work in the industry and domestic work).

***Prison environmental effects (Inputs sheet; optional):*** We include a relative risk multiplier which aims to incorporate additional factors facilitating transmission in prisons in addition to the number of contacts and the underlying per contact transmission probability. By default, we set the environmental relative risk to 1.0 but the user may wish to adjust this to account for specific environmental issues (such as overcrowded cells, shared toilet facilities in cells, or generally unhygienic conditions). Our recommendation is to incorporate these effects through adjustments to contacts and the underlying transmission probability where possible unless there is good evidence to modify from the default value.

**COVID-19 Epidemic Initiation (*Inputs* sheet):**

***Day infected people enter prison*:** The potential day that an infected person enters the prison (Day 0 to Day 120) to begin an epidemic in the setting.

***Mode of entry of infected population:*** The number of infected people coming into the prison via different routes (inmate through reception, inmate through another centre, correctional/healthcare staff, non-essential staff, and family visitors) on the day of entry specified. For family visitors, this is specified as the average number of infected people entering the prison each day from the day of entry specified.

***Variables* sheet inputs:**

This section summarises the model input parameters on the *Variables* sheet. Sources, references, and justifications for each parameter are provided on the *Variables* sheet.

***Age-dependent variables:*** Probability of showing symptoms, probability of hospitalization, probability of going to ICU among hospitalized, proportions of vulnerable, and infection fatality rates per age group (adjusted for vulnerability). Note the model assumes deaths only occur in the hospitalized population and the infection fatality weights are adjusted accordingly within the model.

***Transmission probability per contact:*** Probability of SARS-CoV-2 infection per contact with an infected person (estimated using a transmission model with heterogeneous contact rates with data from Wuhan City, China. London School of Hygiene & Tropical Medicine).

***Relative transmission probability depending on the population or health status***: The health status of the COVID-19 infected status is important for determining the transmission probability, based on disease stage, average relative transmission probabilities are provided (Infectious stage=1 as a baseline, 0 for Exposure and 0.2 for Moderate/Severe/Hospitalized). Health care staff are also at a higher risk of being infected.

***Disease progression rate***: The average rate that individuals to progress through each stage of COVID-19 are specified by the average time in each compartment.

**Effectiveness of Interventions**

These inputs describe the effects of interventions put in place to prevent COVID-19 transmission. This includes the Protective effectiveness of Personal Protective Equipment (PPE) defined by the percentage of reduction in SARS-CoV-2 transmission probability per contact due to handwashing, wearing gloves, and masks in the setting; the effectiveness of thermal scanner in detecting the high temperature of people entering the prison with COVID-19; and the length of quarantine and isolation periods.

**3. Setting up scenarios**

The model can run two outbreak scenarios simultaneously (Scenario 1 and Scenario 2). These scenarios are set-up at the bottom of the *Inputs* sheet. Intervention inputs (shaded in yellow) can be specified to assess the potential impact of interventions on COVID-19 transmission. Through appropriate inputs, the user can run a counterfactual scenario (where inputs are set to the values prior to the COVID-19 pandemic) versus a combination of intervention strategies or compare two specific intervention strategies. Each intervention is described separately below. Specific interventions can be turned on and off using the toggle cells shaded in blue.

**Intervention 0: Reducing the prisoner population**

This intervention is to assess the impact of changes in population size on a COVID-19 outbreak. It is specified by entering the number of inmates who enter and leave the prison setting each day. The prison setting population can only fall to the specified minimum population. Inmates can either enter from the community or via transfer from another correctional centre. If the prison setting has a reception, then inmates will enter the general prison setting population unless the reception quarantine intervention is turned on (see Intervention 3 below). Inmates transferring from another centre are assumed to have already undergone quarantine in another centre and hence join the general inmate population. Only individuals who are susceptible to having mild symptom are assumed to leave prison. Individuals are assumed to stay and isolated in prison once they develop moderate/severe disease and are hospitalized.

**Intervention 1: Controlling the number of contacts each day**

This intervention described the potential control of the number of contacts per inmate per day potentially through social distancing measures, reduction in group sizes outside of their cell, or reductions in time out of the cell.

**Intervention 2: PPE intervention (handwashing, globes, masks)**

This intervention describes the use of PPE in the prison setting to prevent the spread of infection. The user can control the level of PPE intervention throughout the centre by selecting options for ‘only apply to symptomatic inmates’, ‘supply to all inmates’, and ‘supply to staff’. Healthcare staff are assumed to have adequate PPE.

**Intervention 3: Quarantine of incoming inmates in reception**

This intervention described the quarantining of inmates in reception before coming into the prison. Due to logistical constraints, incoming inmates may only be quarantined within a cohort rather than individually. The inputs specify the size of these cohorts and the maximum number of inmates that can be quarantined. The transmission probability of SARS-CoV-2 per contact for people in quarantine is adjusted so that the average number of infections due to an infected inmate during quarantine equals the cohort group size. If the maximum capacity of quarantine in the reception is reached, then inmates will directly go into the prison without quarantine.

**Intervention 4: Isolating inmates with suspected or proven infection**

This intervention describes the isolation of inmates within the general prison to prevent a COVID-19 outbreak. As for intervention 3, inmates may be isolated within a cohort and the transmission probability is adjusted in this setting so the average number of infections within the isolated population equals the cohort size. If the maximum capacity of isolation in the centre has reached, the inmates will continue to homogenously mix within the prison setting.

**Intervention 5: Prevent infected staff attending site**

This intervention describes the prevention of correctional/healthcare staff who are suspected of being in the infectious stage or with symptoms from working. We assume staff still come to the site if they are in the susceptible or exposed stages. They stay away until recovered.

**Intervention 6.1/7.1: Thermal screening of non-essential workers/family visitors**

When this intervention is turned on thermal screening of visitors is in place. People detect to have a fever (based on the effectiveness specified on the *Variables* sheet) are prevented from entering the prison setting.

**Intervention 6.2/7.2: Controlling numbers of non-essential workers/ family visitors.**

When this intervention is turned on the number of visitors coming to the site during the day is reduced by the percentage entered by the user.
